## Supporting information for "Incorporating human mobility to enhance epidemic response and estimate real-time reproduction numbers"

### Effect of mobility and regional epidemic condition on disease transmission

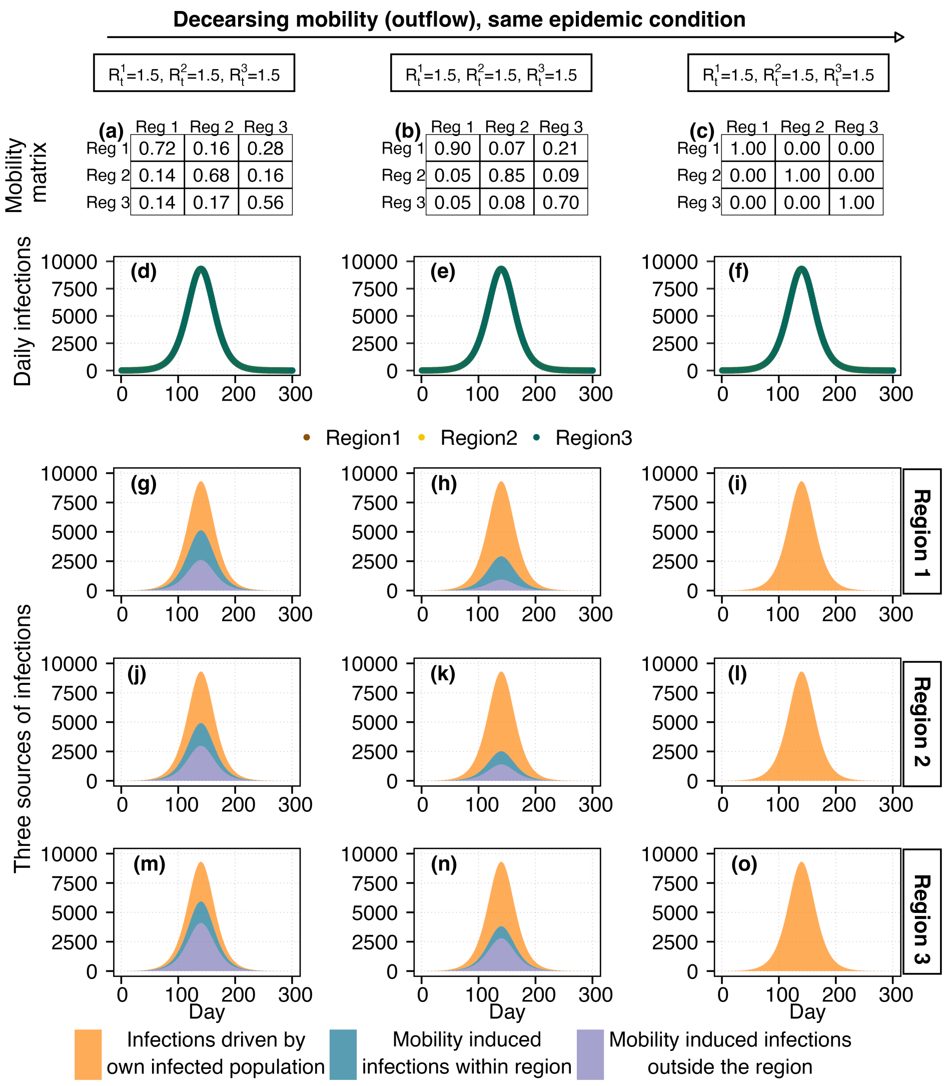


**Fig A: Impact of mobility in a three-region system with identical epidemic conditions.** Each region has an equal population of 1,000,000, the mobility matrices and $R_t$ values are specified in the figure. From left to right, we consider the scenarios with decreasing outgoing mobility and increasing non-commuting population. In the rightmost panel (c) the regions are disconnected (no inter-regional travel), so the mobility matrix is an identity matrix (all off-diagonal elements are zero). Figs. (d)-(f) show daily infections by region. Since the epidemic conditions are homogeneous, the incidence trajectories are identical across regions even when outflows differ. However, mobility redistributes the location of infection acquisition. Figs. (g)-(o) show that, although daily case counts are equal for the three regions, infections occur in different destination regions depending on the mobility pattern.

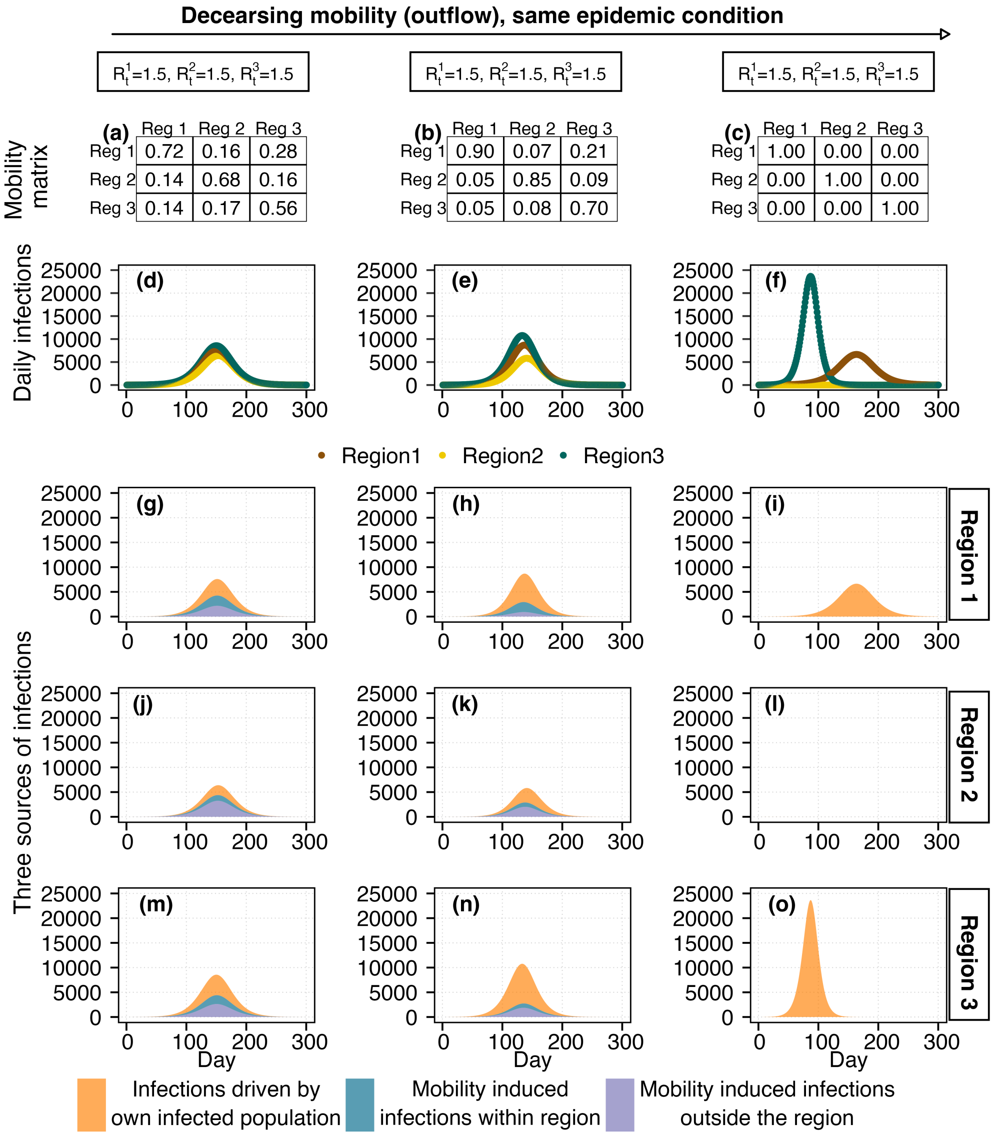


**Fig B:** **Impact of mobility in a three-region system with heterogeneous epidemic conditions.** Each region has an equal population of 1,000,000, the mobility matrices and $R_t$ values are specified in the figure. From left to right, we consider the scenarios with decreasing outgoing mobility and increasing non-commuting population. In the rightmost panel (c) the regions are disconnected (no inter-regional travel), so the mobility matrix is an identity matrix (all off-diagonal elements are zero). Figs. (d)-(f) show daily infections by region. As the epidemic conditions are heterogeneous over regions, the incidence trajectories are different based on the mobility and local epidemic condition. For example, region 2 is with a controlled situation with $R_t < 1$, however, in Figs.(d) and (e), there is an outbreak because of the mobility-driven importation, and in the absence of any mobile population it fails to generate any outbreak (See Fig.(f)). Figs. (g)-(o) show the different sources of infections occur in different destination regions depending on the mobility pattern.

These simulations underscore that both human mobility and regional epidemic status jointly shape disease transmission dynamics. Mobility has a significant impact in heterogeneous epidemic scenarios, where differences in transmission intensity across regions can lead to outbreaks in low-risk areas through population movement. This is particularly relevant in real-world settings, where such heterogeneity is common during the early stages of an epidemic. As the epidemic becomes widespread and more uniform across regions, local disparities can appear due to the differences in policy implementation, healthcare capacity, demographic characteristics, or socio-economic conditions.

**(a)**

| | North East | North West | Yorkshire and the Humber | East Midlands | West Midlands | East | London | South East | South West |
|---|---|---|---|---|---|---|---|---|---|
| South West | 0.00 | 0.00 | 0.00 | 0.00 | 0.00 | 0.00 | 0.00 | 0.01 | 0.97 |
| South East | 0.00 | 0.00 | 0.00 | 0.02 | 0.00 | 0.01 | 0.03 | 0.89 | 0.01 |
| London | 0.00 | 0.00 | 0.00 | 0.01 | 0.00 | 0.04 | 0.94 | 0.09 | 0.00 |
| East | 0.00 | 0.00 | 0.00 | 0.02 | 0.00 | 0.94 | 0.02 | 0.01 | 0.00 |
| West Midlands | 0.00 | 0.01 | 0.00 | 0.03 | 0.98 | 0.00 | 0.00 | 0.00 | 0.00 |
| East Midlands | 0.00 | 0.00 | 0.01 | 0.90 | 0.01 | 0.00 | 0.00 | 0.00 | 0.00 |
| Yorkshire and the Humber | 0.00 | 0.01 | 0.98 | 0.02 | 0.00 | 0.00 | 0.00 | 0.00 | 0.00 |
| North West | 0.00 | 0.97 | 0.00 | 0.01 | 0.00 | 0.00 | 0.00 | 0.00 | 0.00 |
| North East | 0.99 | 0.00 | 0.00 | 0.00 | 0.00 | 0.00 | 0.00 | 0.00 | 0.00 |

**(b)**

| | Hartlepool | Middlesbrough | Redcar and cleveland | Stockton-on-tees | Darlington | County durham | Northumberland | Gateshead | Newcastle upon tyne | North tyneside | South tyneside | Sunderland |
|---|---|---|---|---|---|---|---|---|---|---|---|---|
| Sunderland | 0.01 | 0.00 | 0.00 | 0.00 | 0.00 | 0.04 | 0.01 | 0.04 | 0.01 | 0.01 | 0.07 | 0.86 |
| South tyneside | 0.00 | 0.00 | 0.00 | 0.00 | 0.00 | 0.00 | 0.00 | 0.02 | 0.01 | 0.01 | 0.79 | 0.02 |
| North tyneside | 0.00 | 0.00 | 0.00 | 0.00 | 0.00 | 0.00 | 0.04 | 0.02 | 0.05 | 0.75 | 0.02 | 0.01 |
| Newcastle upon tyne | 0.00 | 0.00 | 0.00 | 0.00 | 0.01 | 0.02 | 0.07 | 0.12 | 0.85 | 0.15 | 0.05 | 0.03 |
| Gateshead | 0.00 | 0.00 | 0.00 | 0.00 | 0.00 | 0.02 | 0.02 | 0.75 | 0.04 | 0.03 | 0.04 | 0.03 |
| Northumberland | 0.00 | 0.00 | 0.00 | 0.00 | 0.00 | 0.01 | 0.85 | 0.02 | 0.03 | 0.04 | 0.01 | 0.00 |
| County durham | 0.03 | 0.01 | 0.00 | 0.02 | 0.06 | 0.86 | 0.01 | 0.03 | 0.01 | 0.01 | 0.01 | 0.04 |
| Darlington | 0.01 | 0.01 | 0.01 | 0.02 | 0.87 | 0.02 | 0.00 | 0.00 | 0.00 | 0.00 | 0.00 | 0.00 |
| Stockton-on-tees | 0.05 | 0.08 | 0.05 | 0.83 | 0.04 | 0.01 | 0.00 | 0.00 | 0.00 | 0.00 | 0.00 | 0.00 |
| Redcar and cleveland | 0.01 | 0.05 | 0.83 | 0.02 | 0.01 | 0.00 | 0.00 | 0.00 | 0.00 | 0.00 | 0.00 | 0.00 |
| Middlesbrough | 0.02 | 0.84 | 0.10 | 0.07 | 0.02 | 0.00 | 0.00 | 0.00 | 0.00 | 0.00 | 0.00 | 0.00 |
| Hartlepool | 0.86 | 0.01 | 0.01 | 0.02 | 0.00 | 0.01 | 0.00 | 0.00 | 0.00 | 0.00 | 0.00 | 0.00 |

**Fig C: Mobility matrix** In this figure, the mobility matrices illustrate the commuting patterns across (a) regions of England and (b) Lower Tier Local Authorities (LTLAs) within the North East region. Each column in the matrix represents the fraction of a region's (or LTLA's) population that commutes to other regions (LTLAs), signifying the outflow from the corresponding region (LTLA) indicated in the figure. On the other hand, the rows depict the inflow to any given region (LTLA) from all other regions (LTLAs). Notably, the diagonal elements (indicated by green box) represent the fraction of the non-commuting population, those who remain within their own region (LTLA). As each column reflects the distribution of a region's (LTLA's) population across all destinations, the sum of each column is equal to one.

In Fig. (a), the mobility data for the regions of England reveals that a significant majority of the population does not commute to other regions on a daily basis for work or other purposes. This may be the reason for the lack of observable impact at this broader spatial resolution. In contrast, Figure (b) details a substantial number of commuters among LTLAs within the North East region of England. The data indicates that approximately 15% to 25% of individuals are commuting to other LTLAs, which appears to significantly influence transmission dynamics.

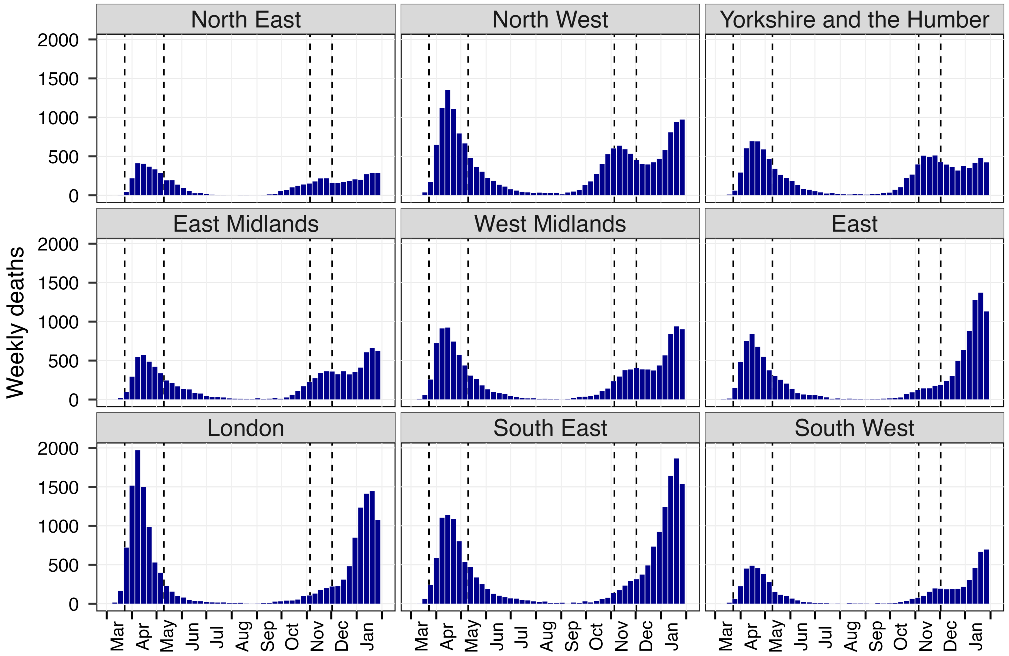


**Fig D: Weekly death data over the nine regions of England.** Vertical lines show the lockdown periods in England.

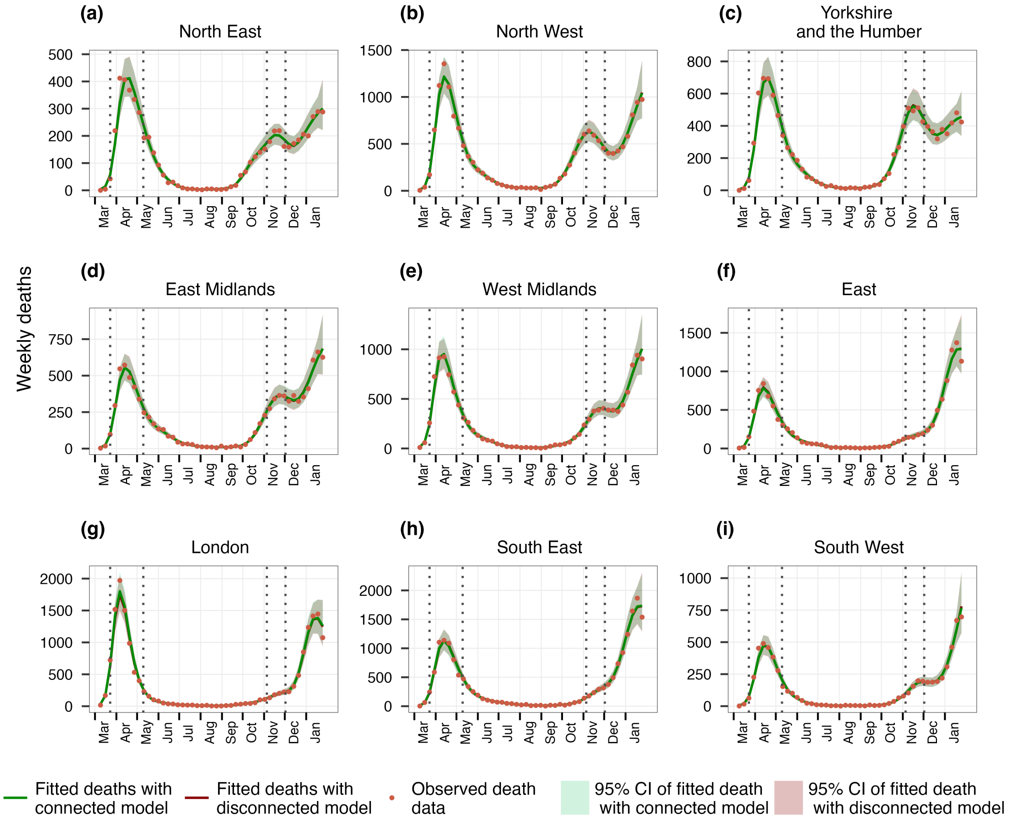


**Fig E: Model Fitting with the connected and disconnected model for nine regions of England.** The green (red) curve is the fitting with the connected (disconnected) model and 95% credible intervals are shown around the curve. Orange dots are the weekly deaths observed for each region. Dotted vertical lines show the lockdown periods.

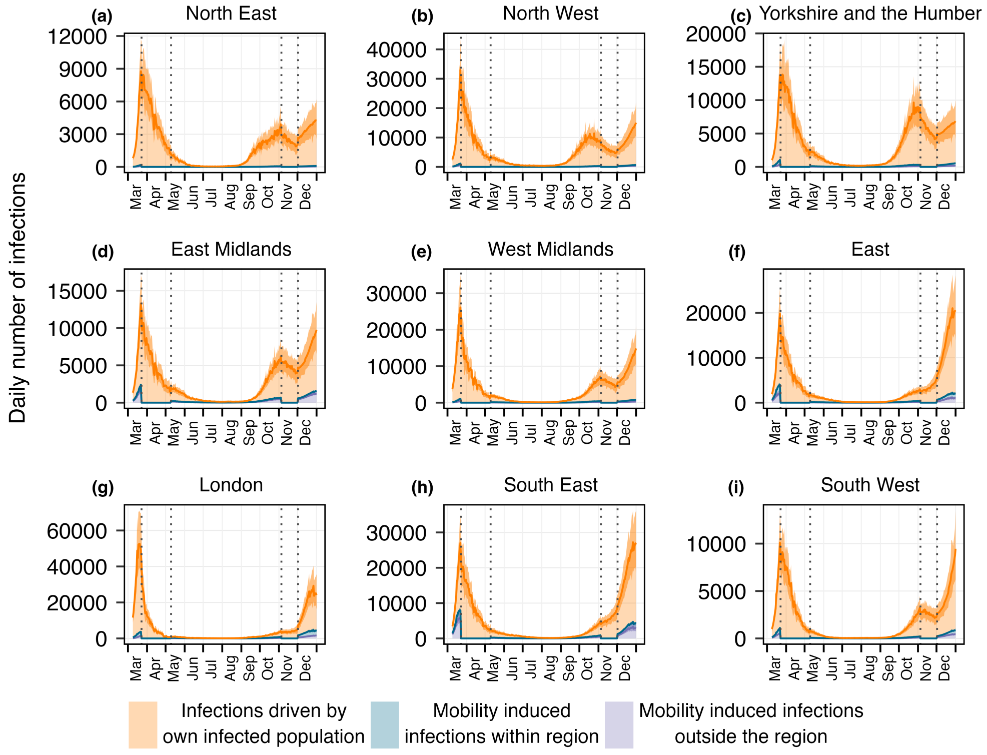


**Fig F: Estimated daily infections across nine regions of England from three different sources.** The colored segments illustrate the contributions from each source: orange indicates infections generated by the own infected population within their own region, blue denotes infections acquired locally due to visits of infected individuals from other regions, and purple represents infections contracted outside the region as a result of travel. Vertical lines show the lockdown periods.

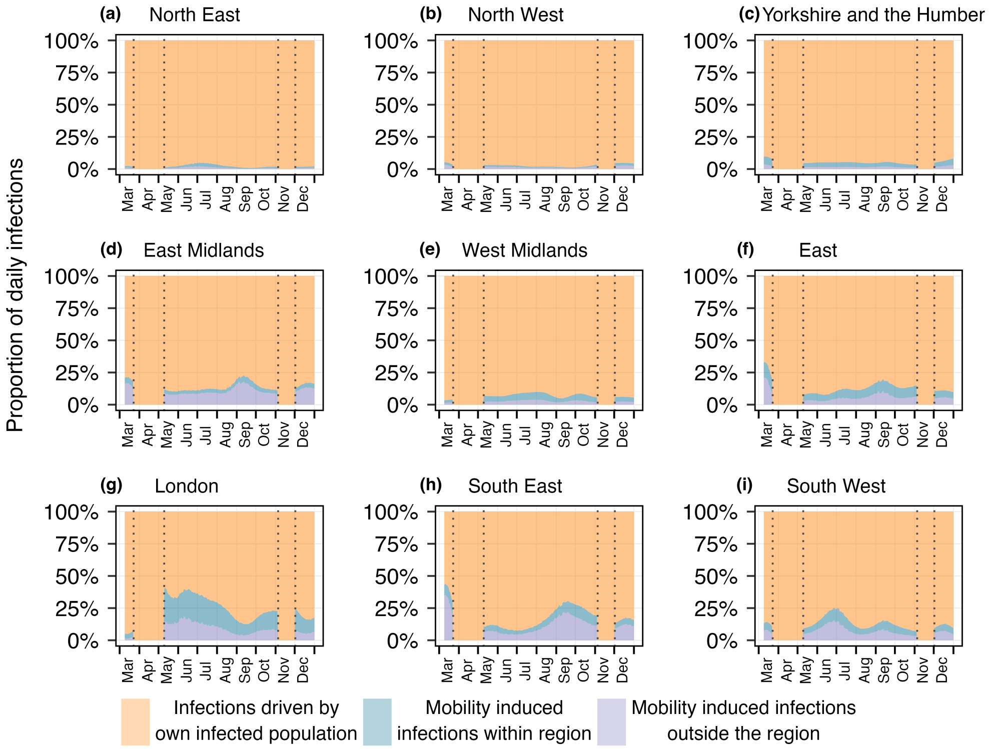


**Fig G: Proportion of daily infections for nine regions of England** The proportion of daily infections coming from three different sources and vertical dotted lines represent the lockdown periods.

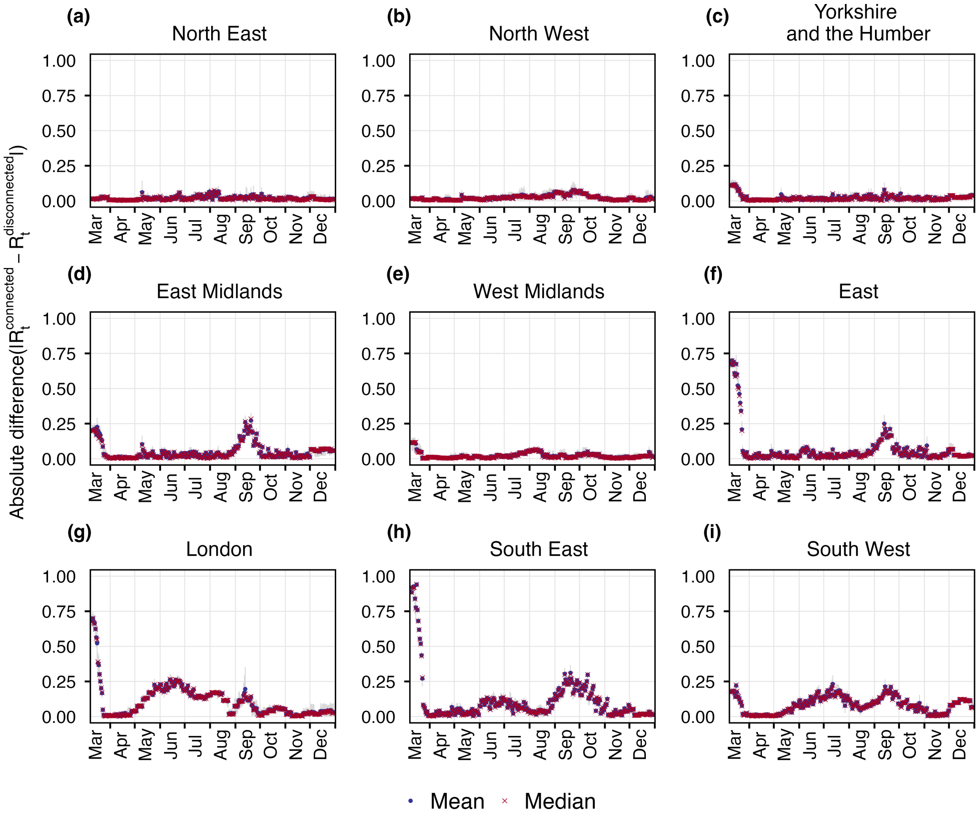


**Fig H: Absolute residuals between connected and disconnected $R_t$s (shown in Fig.4 in the main text) for 9 regions of England.** Blue dots and red crosses show the mean and median of the absolute differences at each time point. Grey bars show the corresponding 95% credible intervals.

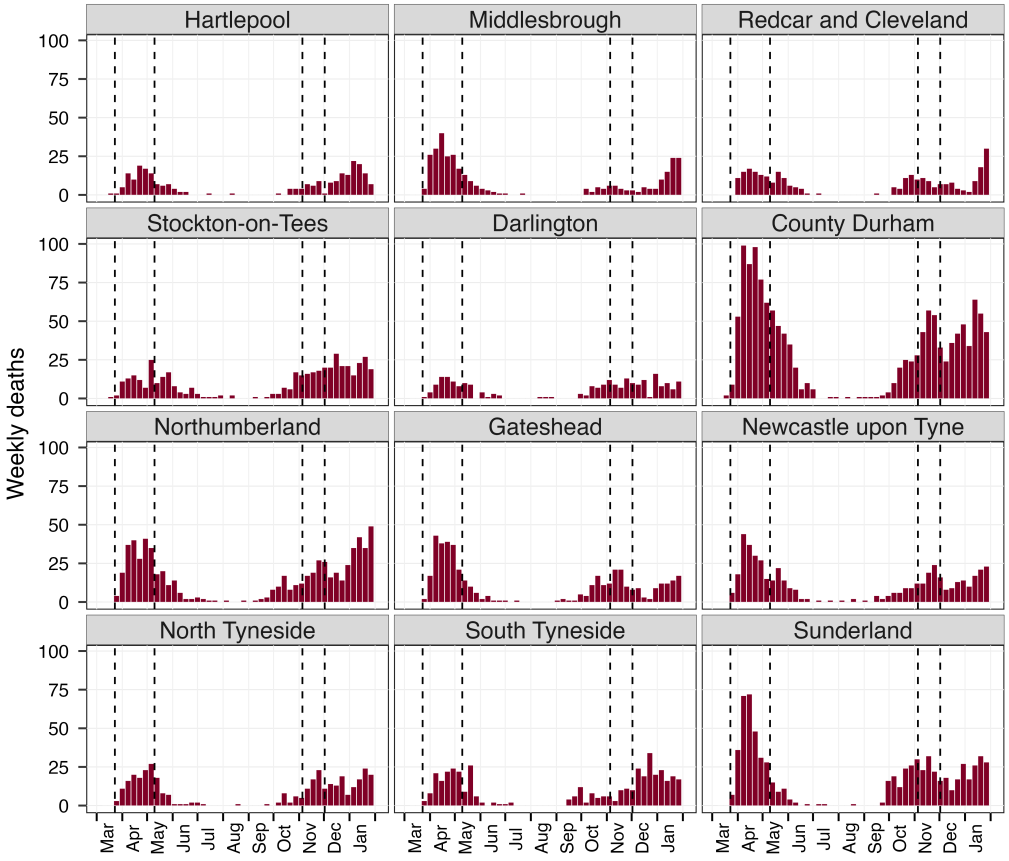


**Fig I: Weekly death data over the twelve LTLAs of North East region.** Vertical lines show the lockdown periods in England.

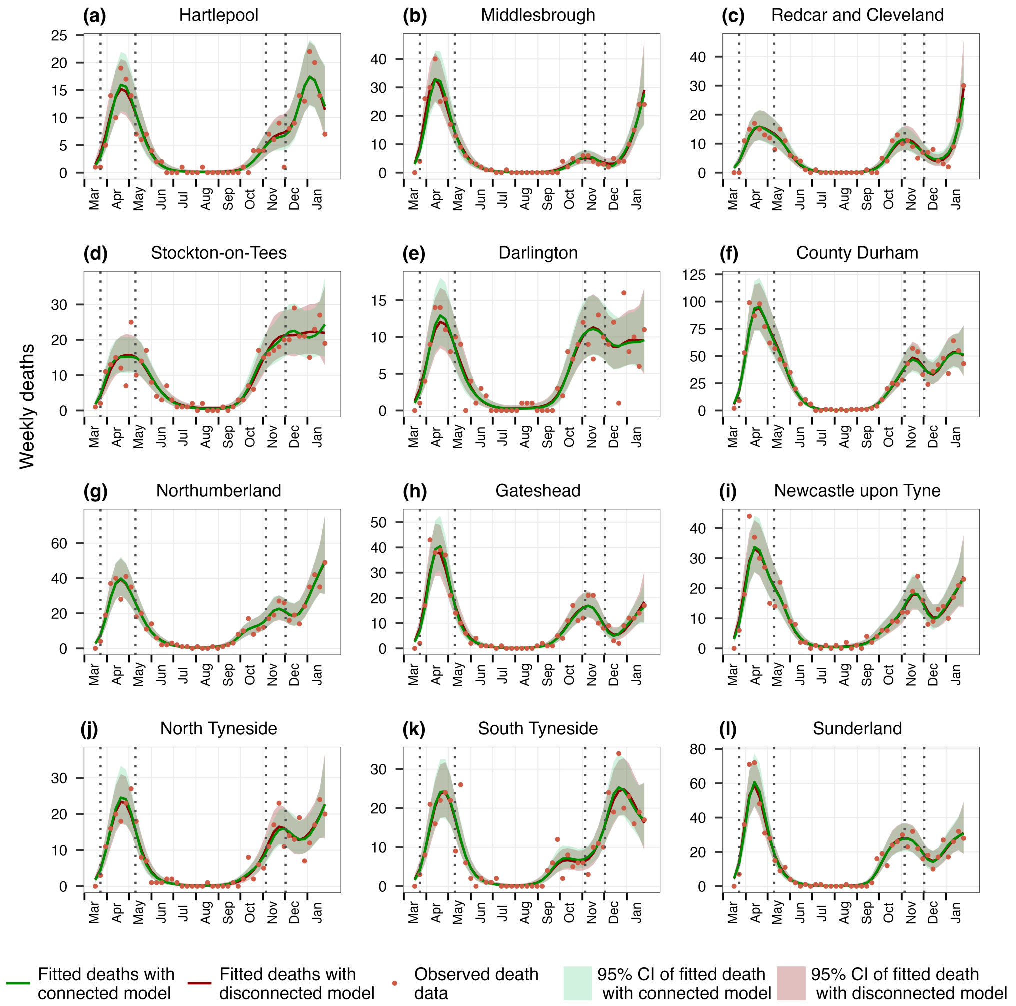


**Fig J: Model Fitting with the connected and disconnected model for twelve LTLA's of North East region.** The green (red) curve is the fitting with the connected (disconnected) model and 95% credible intervals are shown around the curve. Orange dots are weekly deaths observed for each LTLA. Dotted vertical lines show the lockdown periods.

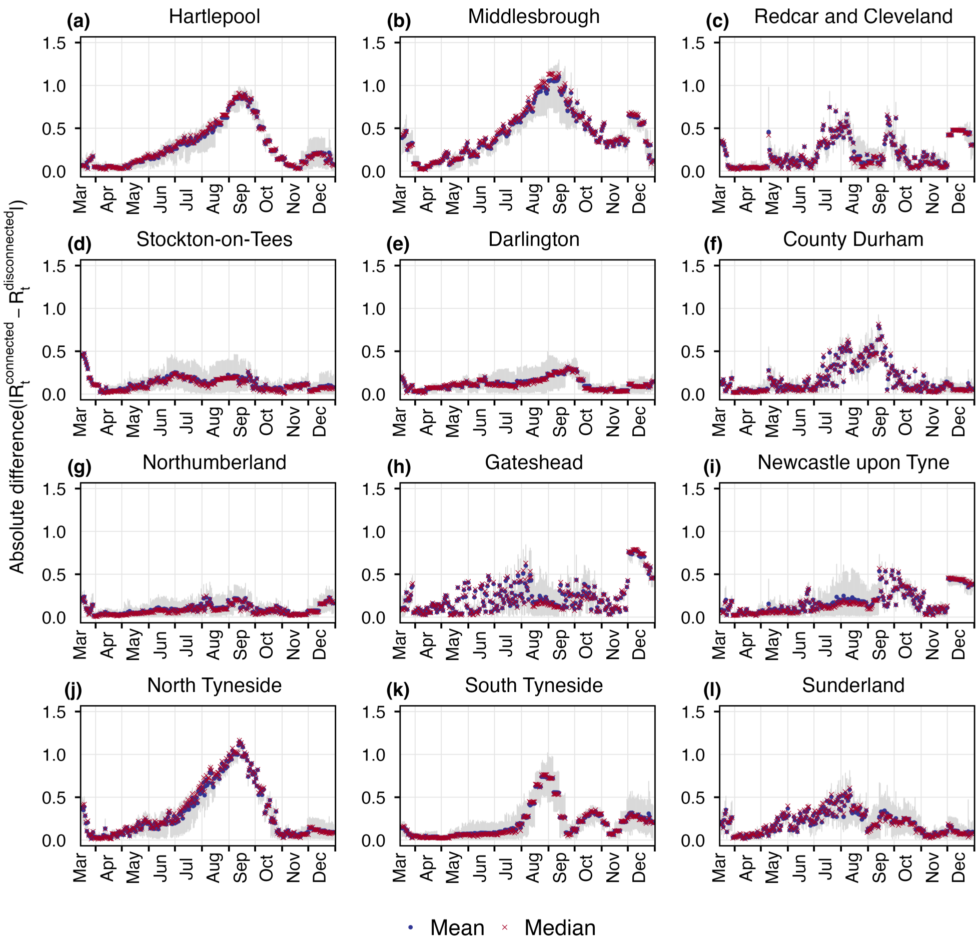


**Fig K: Absolute residuals between connected and disconnected $R_t$s (shown in Fig.5 in the main text) for 12 LTLAs of North East region of England.** Blue dots and red crosses denote the mean and median of the absolute differences at each time point. Grey bars show the corresponding 95% credible intervals.

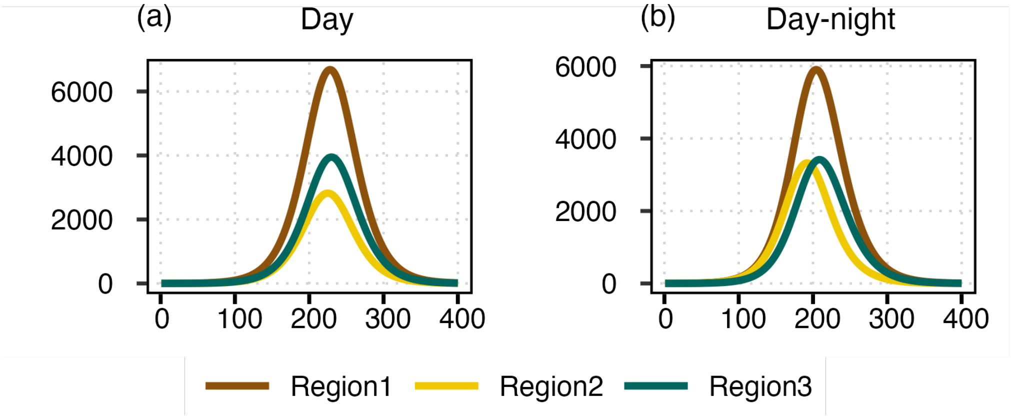


**Fig L: Flexibility of the framework in adapting different time-scales.** We consider three regions with populations of 15,00,000, 5,00,000, 10,00,000, respectively. The reproduction numbers are $R_t^{(1)} = 1.1$, $R_t^{(2)} = 1.7$, $R_t^{(3)} = 1.1$, where $R_t^{(i)}$ represents the reproduction number at time $t$ for region $i$. The mobility matrices are considered as follows:

$$C_{day} = \begin{bmatrix} & \text{Region1} & \text{Region2} & \text{Region3} \\ \text{Region1} & 0.7 & 0.07 & 0.21 \\ \text{Region2} & 0.25 & 0.85 & 0.09 \\ \text{Region3} & 0.05 & 0.08 & 0.7 \end{bmatrix}$$

$$C_{night} = \begin{bmatrix} & \text{Region1} & \text{Region2} & \text{Region3} \\ \text{Region1} & 1 & 0 & 0 \\ \text{Region2} & 0 & 1 & 0 \\ \text{Region3} & 0 & 0 & 1 \end{bmatrix}$$

where, $C_{ij}$ denotes the fraction of population of region $j$ commute to region $i$.

In Fig.(a), we consider a daily time-scale, where the mobility matrix is represented by $C_{day}$ and remains constant throughout the unit time (a full day). In Fig.(b), we consider a finer time scale as half-day intervals. For the first half of the day the mobility matrix is considered as $C_{day}$ and for the second half of the day the mobility matrix is $C_{night}$ to reflect that people typically return to their home regions at night. The generation time distribution is also adjusted accordingly.

Here, $R_t^{(2)} > R_t^{(1)}, R_t^{(3)}$ and therefore, in Fig.(b), when population spends half of the day in their home regions, infections increase in region 2 and decrease in region 1 and region 3 in comparison to Fig.(a).

This shows the flexibility of this framework in adapting different time scales.

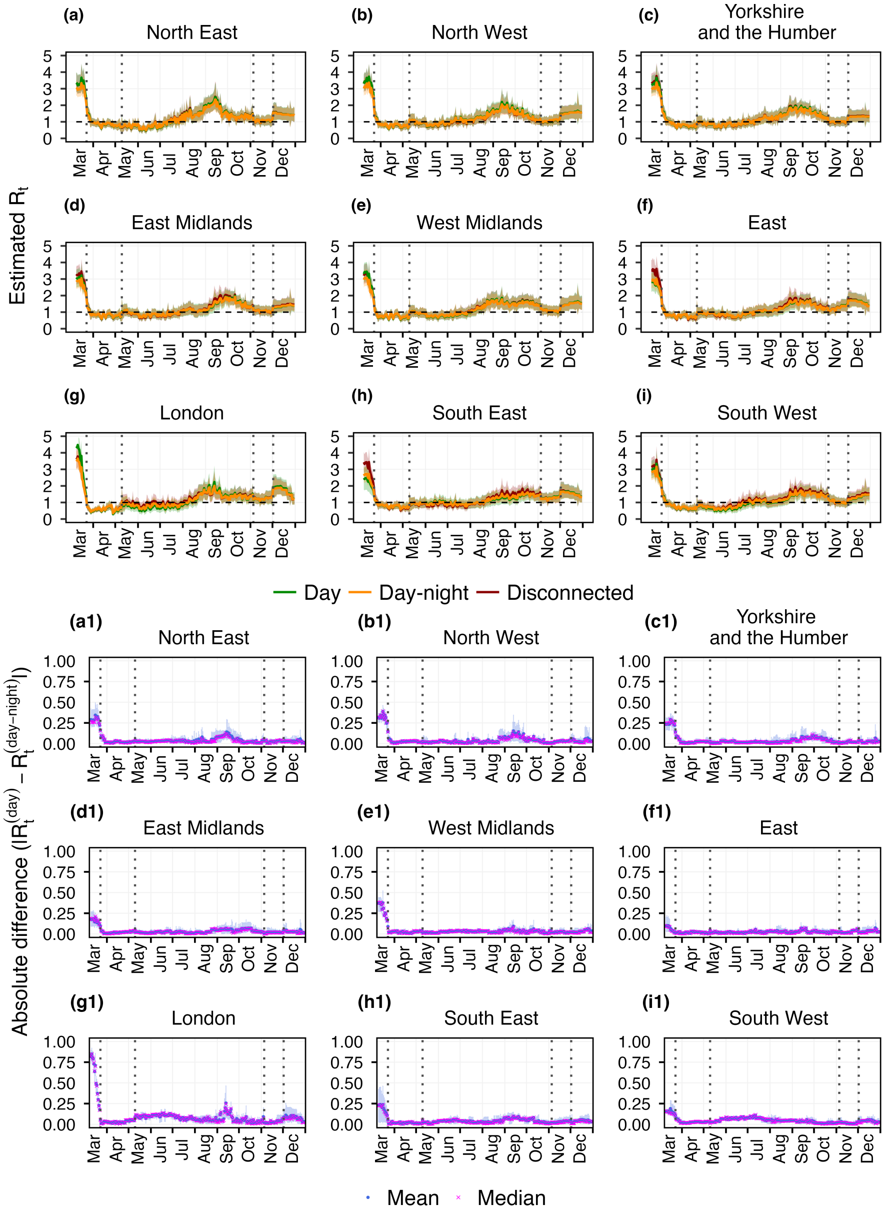


**Fig M: Effect of time scale on the estimated $R_t$ over the regions of England.** We consider two time scales: "day" (same as connected $R_t$ as in Fig.4 in the main text), which are represented by the green curves in (a)-(i), "day-night", where unit time is half of a day and represented by the orange curves, and disconnected $R_t$s are shown by the red curves. In the "day-night" case, the first half of a day uses the mobility matrix as shown in SI Fig B(a), and for the second half we consider the identity matrix to account for the fact that people return to their home region at night. No such difference appears in the results based on the time-scale. We quantify this by calculating absolute residuals $|R_{t,i}^{(day)} - R_{t,i}^{(day-night)}|$ between green and orange curves (see Figs. (a1)-(i1)). Here, blue dots and purple crosses are the mean and median of the residual distribution at each time point, respectively. The corresponding error bars are represented in light blue color.

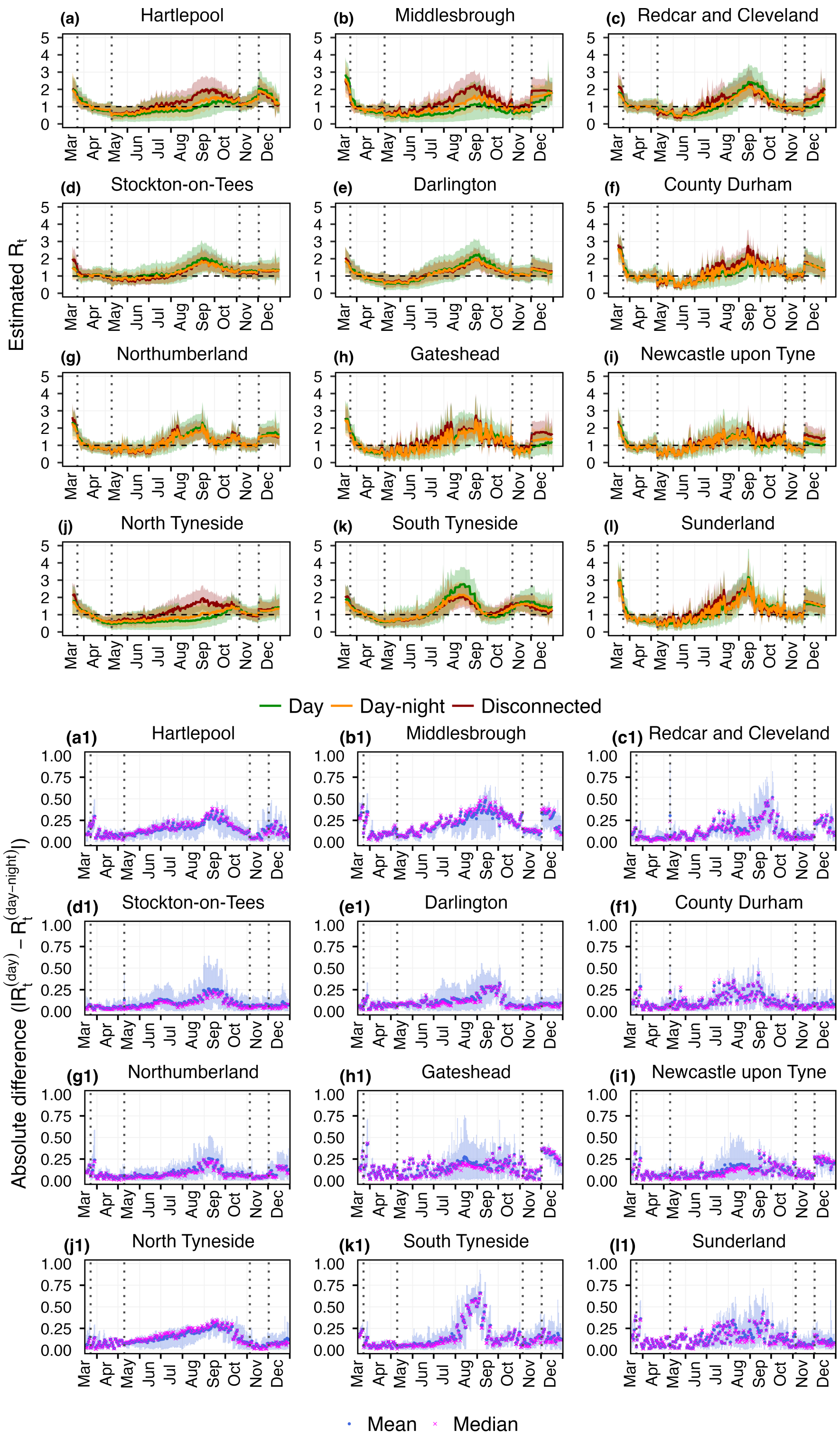


**Fig N: Effect of time scale on the estimated $R_t$ for the LTLAs of North East region.** We consider two time scales: "day" (same as connected $R_t$ as in Fig.5 in the main text), which are represented by the green curves in (a)-(l), "day-night", where unit time is half of a day and represented by the orange curves, and disconnected $R_t$s are shown by the red curves. In the "day-night" case, the first half of a day uses the mobility matrix as shown in SI Fig B(b), and for the second half we consider the identity matrix to account for the fact that people return to their home region at night. We quantify this by calculating absolute residuals $|R_{t,i}^{(day)} - R_{t,i}^{(day-night)}|$ between green and orange curves (see Figs. (a1)-(l1)). Here, blue dots and purple crosses are the mean and median of the residual distribution at each time point, respectively. The corresponding error bars are represented in light blue color. The choice of time scale does not lead to any fundamental change in the obtained results.

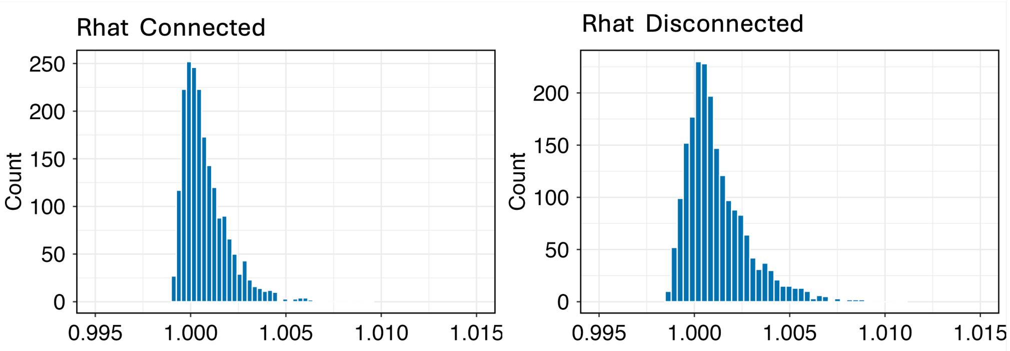


**Fig O: Distribution of Rhat statistics.** Values close to 1 indicate MCMC convergence.
